# Mortality outcomes with hydroxychloroquine and chloroquine in COVID-19: an international collaborative meta-analysis of randomized trials

**DOI:** 10.1101/2020.09.16.20194571

**Authors:** Cathrine Axfors, Andreas M. Schmitt, Perrine Janiaud, Janneke van ’t Hooft, Sherief Abd-Elsalam, Ehab F. Abdo, Benjamin S. Abella, Javed Akram, Ravi K. Amaravadi, Derek C. Angus, Yaseen M. Arabi, Shehnoor Azhar, Lindsey R. Baden, Arthur W. Baker, Leila Belkhir, Thomas Benfield, Marvin A.H. Berrevoets, Cheng-Pin Chen, Tsung-Chia Chen, Shu-Hsing Cheng, Chien-Yu Cheng, Wei-Sheng Chung, Yehuda Z. Cohen, Lisa N. Cowan, Olav Dalgard, Fernando F. de Almeida e Val, Marcus V.G. de Lacerda, Gisely C. de Melo, Lennie Derde, Vincent Dubee, Anissa Elfakir, Anthony C. Gordon, Carmen M. Hernandez-Cardenas, Thomas Hills, Andy I.M. Hoepelman, Yi-Wen Huang, Bruno Igau, Ronghua Jin, Felipe Jurado-Camacho, Khalid S. Khan, Peter G Kremsner, Benno Kreuels, Cheng-Yu Kuo, Thuy Le, Yi-Chun Lin, Wu-Pu Lin, Tse-Hung Lin, Magnus Nakrem Lyngbakken, Colin McArthur, Bryan J. McVerry, Patricia Meza-Meneses, Wuelton M. Monteiro, Susan C. Morpeth, Ahmad Mourad, Mark J. Mulligan, Srinivas Murthy, Susanna Naggie, Shanti Narayanasamy, Alistair Nichol, Lewis A. Novack, Sean M. O’Brien, Nwora Lance Okeke, Léna Perez, Rogelio Perez-Padilla, Laurent Perrin, Arantxa Remigio-Luna, Norma E. Rivera-Martinez, Frank W. Rockhold, Sebastian Rodriguez-Llamazares, Robert Rolfe, Rossana Rosa, Helge Røsjø, Vanderson S. Sampaio, Todd B. Seto, Muhammad Shehzad, Shaimaa Soliman, Jason E. Stout, Ireri Thirion-Romero, Andrea B. Troxel, Ting-Yu Tseng, Nicholas A. Turner, Robert J. Ulrich, Stephen R. Walsh, Steve A. Webb, Jesper M. Weehuizen, Maria Velinova, Hon-Lai Wong, Rebekah Wrenn, Fernando G. Zampieri, Wu Zhong, David Moher, Steven N. Goodman, John P.A. Ioannidis, Lars G. Hemkens

## Abstract

Substantial COVID-19 research investment has been allocated to randomized clinical trials (RCTs) on hydroxychloroquine/chloroquine, which currently face recruitment challenges or early discontinuation. We aimed to estimate the effects of hydroxychloroquine and chloroquine on survival in COVID-19 from all currently available RCT evidence, published and unpublished. We conducted a rapid meta-analysis of ongoing, completed, or discontinued RCTs on hydroxychloroquine or chloroquine treatment for any COVID-19 patients (protocol: https://osf.io/QESV4/). We systematically identified unpublished RCTs (ClinicalTrials.gov, WHO International Clinical Trials Registry Platform, Cochrane COVID-registry up to June 11, 2020), and published RCTs (PubMed, medRxiv and bioRxiv up to October 16, 2020). All-cause mortality was extracted (publications/preprints) or requested from investigators and combined in random-effects meta-analyses, calculating odds ratios (ORs) with 95% confidence intervals (CIs), separately for hydroxychloroquine and chloroquine. Prespecified subgroup analyses included patient setting, diagnostic confirmation, control type, and publication status. Sixty-three trials were potentially eligible. We included 14 unpublished trials (1308 patients) and 14 publications/preprints (9011 patients). Results for hydroxychloroquine are dominated by RECOVERY and WHO SOLIDARITY, two highly pragmatic trials, which employed relatively high doses and included 4716 and 1853 patients, respectively (67% of the total sample size). The combined OR on all-cause mortality for hydroxychloroquine was 1.11 (95% CI: 1.02, 1.20; I^2^=0%; 26 trials; 10,012 patients) and for chloroquine 1.77 (95%CI: 0.15, 21.13, I^2^=0%; 4 trials; 307 patients). We identified no subgroup effects. We found that treatment with hydroxychloroquine was associated with increased mortality in COVID-19 patients, and there was no benefit of chloroquine. Findings have unclear generalizability to outpatients, children, pregnant women, and people with comorbidities.

## Introduction

Coronavirus disease 2019 (COVID-19) caused by severe acute respiratory syndrome coronavirus 2 (SARS-CoV-2) has the potential of progression into respiratory failure and death.^1^ More than 900,000 persons with COVID-19 globally have died by September, 2020,^2^ and treatment options are limited.^3^ The COVID-19 pandemic has caused a hitherto unprecedented search for possible therapies, with almost 700 clinical trials initiated in the first quarter of 2020 - and one in five of these trials target hydroxychloroquine (HCQ) or chloroquine (CQ).^4^ This remarkable attention was primarily due to *in vitro* data,^5^ immunomodulatory capacities,^6^ and the oral formulation and well-documented safety profiles.

In March 2020, the US Food and Drug Administration (FDA) issued an Emergency Use Authorization of HCQ^7^ and its prescription and usage outside clinical studies skyrocketed.^8^ In many countries, HCQ or CQ were listed in treatment guidelines for COVID-19 (including, e.g., China, Ireland, and the US).^9^ In a New York City cohort of 1376 COVID-19 inpatients during March-April 2020, 59% received HCQ.^10^ However, the FDA revoked the Emergency Use Authorization on June 15, 2020.^11^ At that point, two large randomized clinical trials (RCTs), RECOVERY and the WHO Solidarity trial, had stopped enrollment to their HCQ treatment arms.^12,13^ An interim analysis of the RECOVERY trial showed no mortality benefit of HCQ.^13^ Established as treatments of malaria and rheumatic disorders, HCQ and CQ may carry potentially severe adverse effects, especially related to cardiac arrhythmia.^6^ Public uncertainty still remains, as illustrated by recent reports of planned use in pandemic epicenters in Central and South America.^14^

While many trials are ongoing, additional published evidence of potential benefits or harms may be several months away, if they even reach completion. Given the lack of favorable results in the large RECOVERY trial and the revoked Emergency Use Authorization, recruitment into HCQ and CQ trials has become increasingly difficult and many trials may run the risk of ending in futility. A rapid examination of data on all-cause mortality from as many trials as possible may offer the best evidence on potential survival benefits and to ensure that patients are not exposed to unnecessary risks if benefit is lacking. We used the infrastructure established with COVID-evidence,^15^ a comprehensive database of COVID-19 trials funded by the Swiss National Science Foundation, to invite all investigators of HCQ or CQ trials to participate in an international collaborative meta-analysis. We aimed to identify and combine all RCTs investigating the effects of HCQ or CQ on all-cause mortality in patients with COVID-19 compared to any control arm similar to the experimental arm in all aspects except the administration of HCQ or CQ.

## Methods

This collaborative meta-analysis focused solely on all-cause mortality in order to provide rapid evidence on the most critical clinical outcome. Investigators of ongoing, discontinued or completed trials were contacted via email to provide group-level (aggregated) mortality data per trial arm at any time point available. The protocol was published online before data collection.^16^

We considered all clinical trials that reported randomly allocating patients with confirmed or suspected SARS-CoV-2 infection to a treatment protocol containing HCQ or CQ (for any duration or dose) or the same treatment protocol not containing HCQ or CQ. In other words, the control group had to receive placebo or no treatment other than standard of care (we excluded comparisons of HCQ or CQ against an active treatment, e.g., HCQ versus azithromycin, since active controls were too heterogeneous to pool together and reveal the pure benefits and harms of HCQ or CQ). Eligible ongoing trials had to provide data on all-cause mortality and randomize the first patient before June 1, 2020 (time point selected arbitrarily as we did not expect trials launched later to recruit enough patients to provide relevant additional information). Trials published or posted as preprint were not restricted by date. Prevention trials were not included. We included trials regardless of whether mortality was a primary outcome or not and put no restrictions on trial status, language, geographical region, or healthcare setting.

We searched for eligible trials registered at ClinicalTrials.gov and the WHO International Clinical Trials Registry Platform [ICTRP] by June 11, 2020 (COVID-evidence database).^17^ We additionally searched PubMed and the Cochrane COVID-19 trial registry (covering preprints, trial registries and literature databases) by June 11, 2020, using terms related to HCQ and CQ combined with terms for COVID-19 and a standard RCT filter (Supplement 1).^18^ We updated the literature search on October 16, 2020. Two authors (CA and AMS) independently verified the eligibility criteria (Figure 1) and solved any discrepancies by discussion.

**Figure 1.**
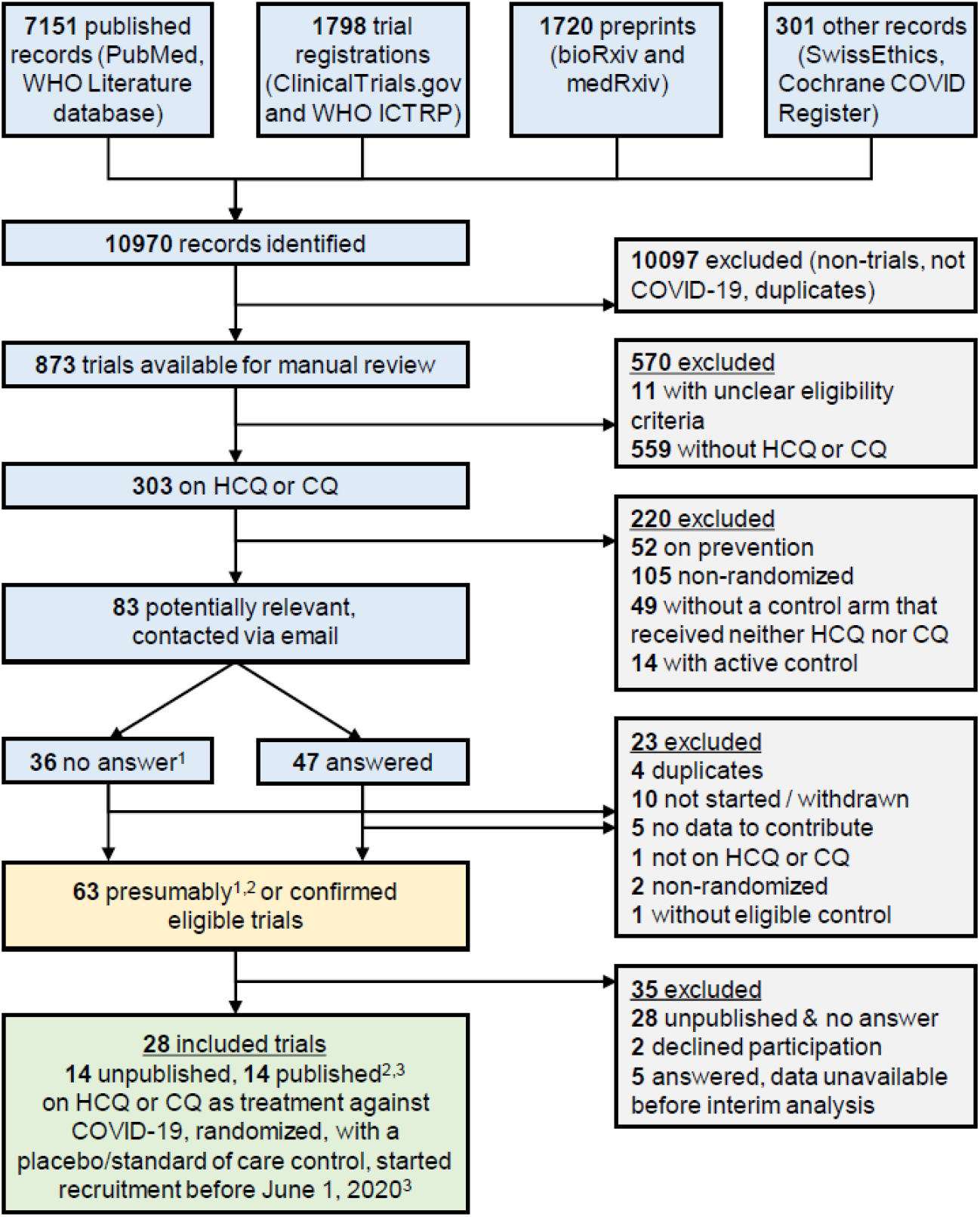
Flowchart of included randomized clinical trials. Sources searched up to June 11, 2020 (PubMed, ClinicalTrials.gov, WHO ICTRP, Cochrane COVID Register) or April 9 (WHO Literature database, bioRxiv, medRxiv, SwissEthics). ^1^ Trials for which we received no answer were presumed to be eligible unless withdrawn. ^2^ One publication and two preprints were identified in a later search update. ^3^ Published peer-reviewed articles or posted preprints. Abbreviations: chloroquine (CQ), hydroxychloroquine (HCQ), International Clinical Trials Registry Platform (ICTRP), World Health Organization (WHO).

Principal investigators of 83 potentially eligible trials were asked to confirm the eligibility criteria, as well as: “For each of your study arms: (a) What intervention did this group receive? (b) How many patients were randomized to this group? (c) Of these patients, how many have died? (d) Of these patients, for how many it is unknown if they are dead or alive?” (Supplement 2, email template). Investigators who were not responsive received two email reminders in English or Chinese, depending on trial origin.

The following information was extracted from all included RCTs by two reviewers (CA, AMS) and verified by the trial investigators: experimental and control arms, number of randomized participants, treatment schedule, patient setting, eligibility criteria, study location, blinding, target sample size, and trial status. We also classified trials as published in a peer-reviewed journal, posted on a preprint server, or unpublished (the latter category not including preprints). For reasons of feasibility within this rapid assessment, we generally did not request descriptive information beyond items included in trial registrations.

The main analysis evaluated separately the effect on all-cause mortality of HCQ versus control and CQ versus control. We report absolute numbers and proportions, as well as the treatment effect estimate as an odds ratio (OR; odds of death in the HCQ or CQ intervention group divided by the odds of death in the control group) with 95% confidence intervals (CIs). For multi-arm studies, we requested data for all arms and calculated treatment effect estimates for each eligible comparison. We combined mortality effects from all RCTs based on binary outcome data (2×2 contingency tables) in meta-analyses and describe the statistical heterogeneity using the I^2^-statistic.^19^In our protocol, we prespecified a random-effects model of the Hartung-Knapp-Sidik-Jonkman (HKSJ) approach,^20^ in order to provide more equality of weights between trials with moderate to large size (than for example the DerSimonian-Laird approach). We did not prespecify the between-study variance estimator, tau-squared, but chose the Paule and Mandel (PM) estimator based on provided guidance on choosing among 16 variants.^21^ Cases of zero events in one arm were corrected by adding the reciprocal of the size of the contrasting study arm.^18^ However, considering the range of sample sizes and numbers of zero events across trials, we assessed the effects of alternative approaches with sensitivity analyses, as detailed below. To explore and illustrate evidence generation over time, we also performed a cumulative meta-analysis of all trials as well as stratified by dissemination status (publications/preprints vs unpublished), using the HKSJ approach with PM tau-squared. We used the date of email response or publication/posting of preprint. The meta-analyses were completed using R version 3.5.1 and the ‘meta ‘package version 4.13-0.

In exploratory subgroup analyses, we stratified trials by patient setting (as proxy to COVID-19 severity: outpatients, inpatients but not intensive care unit (ICU), and ICU), diagnostic confirmation (confirmed SARS-CoV-2 versus suspected cases), control type (placebo control versus other) and publications/preprints versus unpublished trials. We did not stratify for missing data since the amount was extremely low. A post-hoc stratification by HCQ dose was added (trials with ≥1600 mg on day 1 and ≥800 mg from day 2 versus lower-dose trials) to isolate trials predicted to achieve blood levels of HCQ above the *in vitro* half maximal inhibitory concentration (IC50) value for SARS-CoV-2 (1.13 μM).^22^

We added exploratory sensitivity analyses to assess robustness across meta-analytic approaches: DerSimonian-Laird and Sidik-Jonkman tau-squared estimators, Mantel-Haenszel random-effects method, and Peto method. DerSimonian-Laird is a standard random-effects meta-analysis approach, but may underestimate uncertainty. The Sidik-Jonkman tau-squared estimator, on the other hand, may yield inflated estimates if heterogeneity is low.^21^ The Mantel-Haenszel method performs reasonably well with small and zero event counts, much like Peto and arcsine transformation. The Peto method is suboptimal in the presence of substantial imbalances in the allocation ratio of patients randomized in the compared arms (e.g., RECOVERY trial). We also modeled variants to handling zero events (arcsine difference, and excluding trials with zero events) as well as excluding trials with <50 participants.

## Results

Our search identified 146 randomized trials investigating HCQ or CQ as treatment for COVID-19, of which 83 were deemed potentially eligible after scrutinizing the randomized comparisons. The investigators of these 83 trials were contacted and 57% (47 of 83) responded (Figure 1). Of the responders, 19 trials were eligible and available (14 unpublished, three preprints and two publications); 21 trials were ineligible according to information provided; five responding investigator teams were not ready to share their results yet; and two declined participation. For the 36 trials without response, six were confirmed eligible and available (four publications and two preprints); two were confirmed ineligible; and for the remaining 28, results were not available, nor could they be confirmed eligible.

We included 28 trials (14 unpublished trials, eight publications, and six preprints; of these, one publication and two preprints were identified for the first time in our search update).^13,23–35^Individual trial characteristics are presented in Table 1 (28 included trials) and Supplement Table S1 (35 potentially eligible but unavailable). Overall, trial characteristics were not different between included and unavailable trials (Table 2).

**Table 1.**
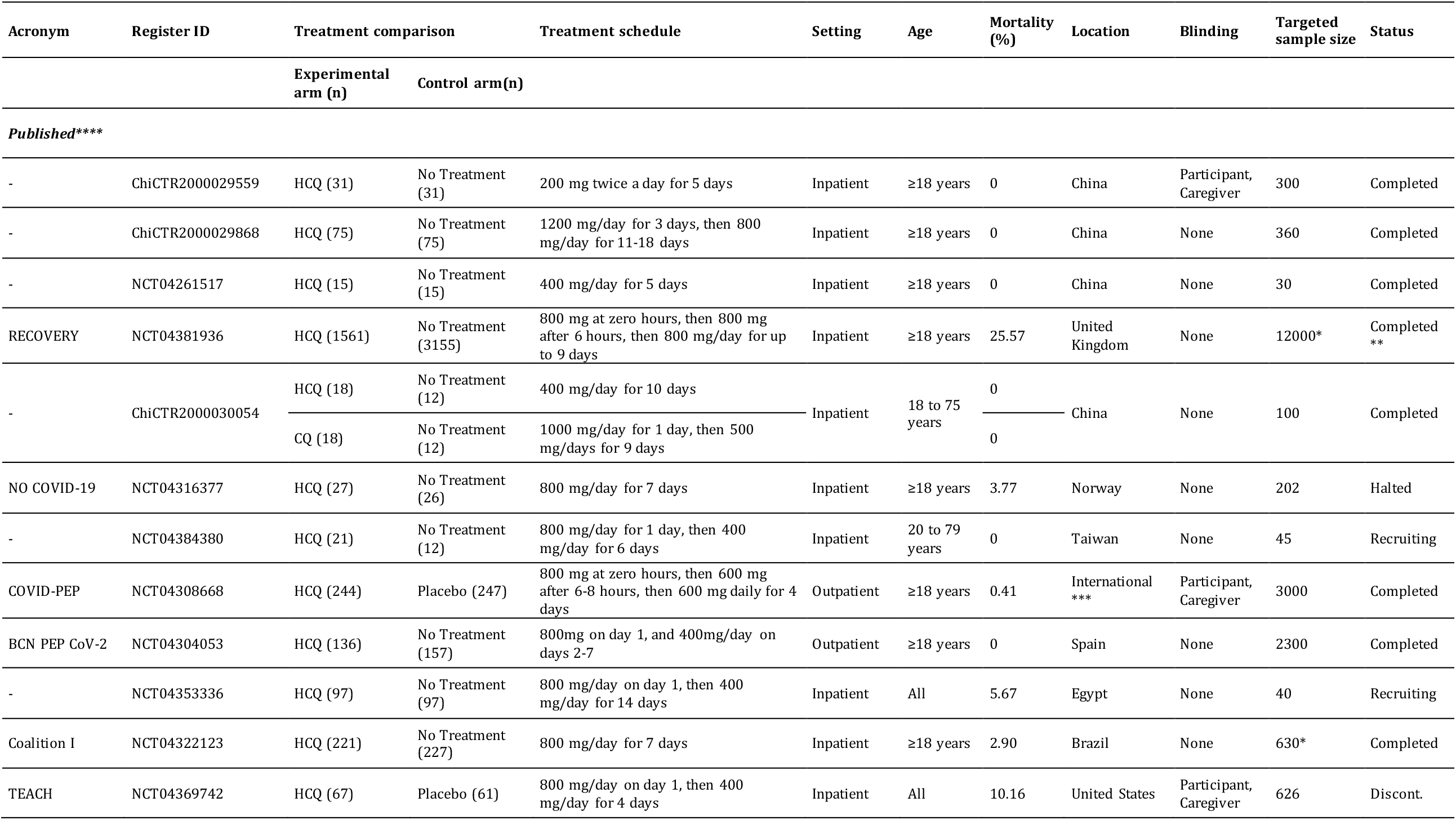
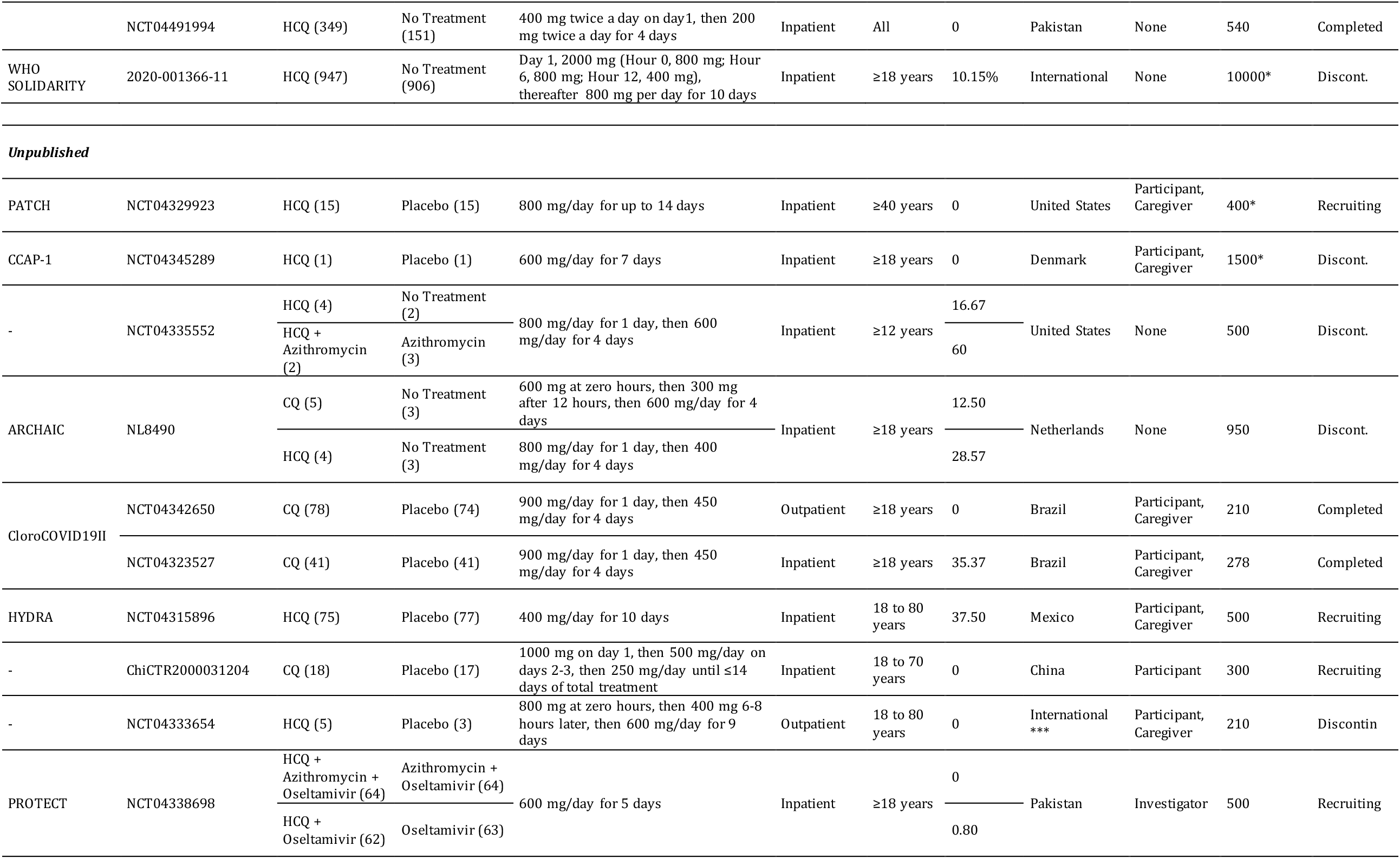
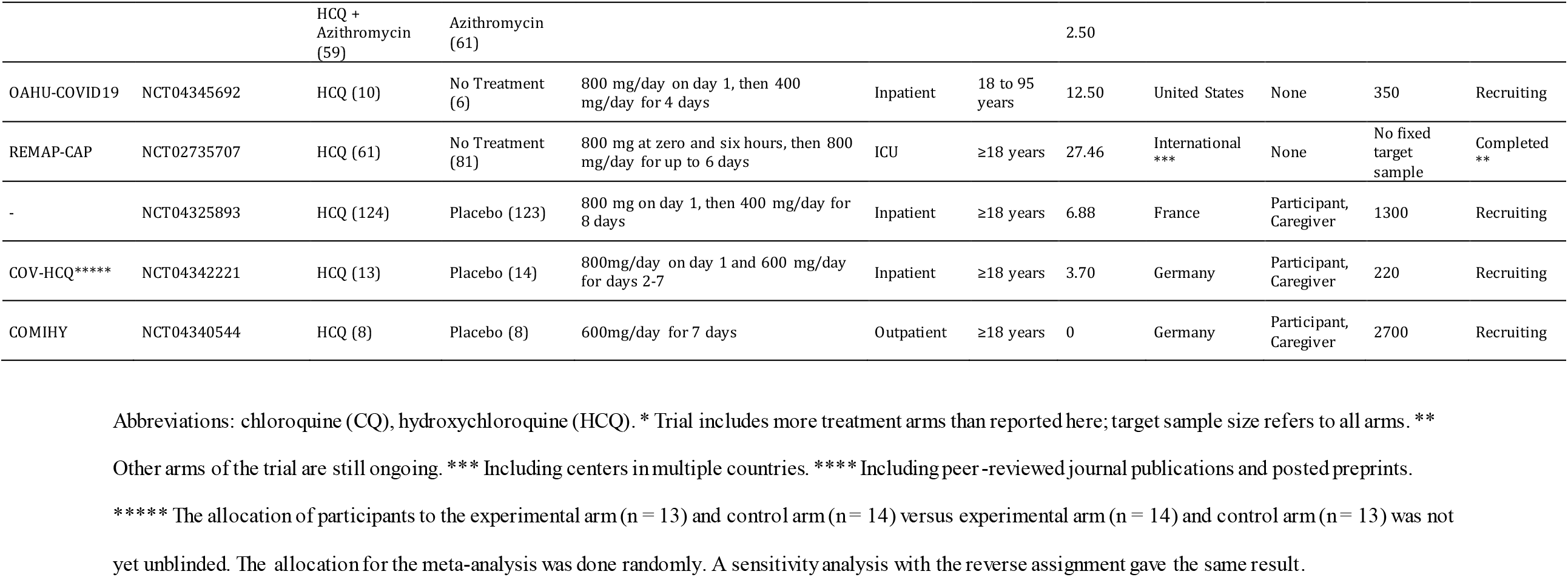
Group-level characteristics of randomized clinical trials evaluating hydroxychloroquine or chloroquine as treatment for COVID-19.

**Table 2.** Group-level characteristics of included and unavailable trials.

|  | **All trials**<br>**n = 63** | **Included trials**<br>**n = 28** | **Potentially eligible,**<br>**unavailable trials\***<br>**n = 35** |
| --- | --- | --- | --- |
| **Drug, n (%)** |  |  |  |
| HCQ | 48 (76) | 24 (86) | 24 (69) |
| CQ | 10 (16) | 2 (7) | 8 (22) |
| Both | 5 (8) | 2 (7) | 3 (8) |
| **Planned sample size\*, median (IQR)** | 360 (150 to 672) | 500 (218 to 1350) | 267 (120 to 483) |
| **Trial status, n (%)** |  |  |  |
| Completed | 12 (19) | 11 (39) | 1 (3) |
| Discontinued | 15 (24) | 7 (25) | 8 (23) |
| Not yet recruiting | 7 (11) | 0 | 7 (20) |
| Recruiting | 29 (46) | 10 (36) | 19 (54) |
| **Location, n (%)** |  |  |  |
| Africa | 3 (5) | 1 (4) | 2 (6) |
| Asia | 23 (37) | 8 (29) | 15 (43) |
| Europe | 17 (27) | 8 (29) | 9 (26) |
| International | 6 (10) | 4 (14) | 2 (6) |
| North America | 10 (16) | 4 (14) | 6 (17) |
| South America | 4 (6) | 3 (11) | 1 (3) |
| **Placebo control, n (%)** | 30 (48) | 11 (39) | 19 (54) |
| **More than two arms, n (%)** | 27 (44) | 10 (37) | 17 (49) |
| **Patient setting, n (%)** |  |  |  |
| ICU | 1 (2) | 1 (4) | 0 |
| Inpatient | 46 (73) | 22 (79) | 24 (69) |
| Outpatient | 12 (19) | 5 (18) | 7 (20) |
| Unclear | 4 (6) | 0 | 4 (11) |
| **Blinding, n (%)** |  |  |  |
| None | 32 (51) | 15 (54) | 17 (49) |
| Outcome Assessor | 1 (2) | 1 (4) | 0 |
| Participant | 3 (5) | 1 (4) | 2 (6) |
| Participant, Caregiver | 26 (41) | 11 (39) | 15 (43) |
| Participant, Outcome Assessor | 1 (2) | 0 | 1 (3) |

Abbreviations: chloroquine (CQ), hydroxychloroquine (HCQ), intensive care unit (ICU), interquartile range (IQR)
\* Data were extracted from trial registries or publications. \*\* Including centers in multiple countries.

HCQ was evaluated in 26 trials (10,012 patients), and CQ was evaluated in four trials (307 patients). Two trials investigated both HCQ versus control and CQ versus control (63 patients). The median sample size was 95 (IQR 28 to 282) for HCQ trials and 42 (IQR 29 to 95) for CQ trials. The two largest trials (RECOVERY and WHO SOLIDARITY) included 47% and 19% of all patients in the HCQ trials, respectively. Most trials investigated HCQ or CQ in hospitalized patients (22 trials; 79%), and only five trials (18%) had an outpatient setting. The average mortality was 10.3% (standard deviation 13.5%) in inpatient trials and 0.08% (standard deviation 0.18%) in outpatient trials. The comparator was in eleven trials placebo (39%) and in 17 (61%) no other treatment than standard of care. In most trials, patients and clinicians were aware of the treatment (15 trials; 54%), while in one trial (4%) the patients were blinded and in 11 trials (39%) patients and clinicians were blinded (Table 2).

Regarding HCQ, in the 26 included trials, 606 of 4316 (14.0%) patients treated with HCQ died and 960 of 5696 patients (16.9%) in the control groups died (within 19 trials with a 1:1 randomization ratio, 7.7% of patients in the HCQ arm died [181 of 2346] and 7.1% of patients in the control arm died [168 of 2352]). In the meta-analysis, the combined OR was 1.11 (95% CI, 1.02 to 1.20, p = 0.02), with low heterogeneity (I^2^ = 0%) (Figure 2A). In 12 trials including a total of 1282 patients, there were zero deaths in both arms.

**Figure 2A.**
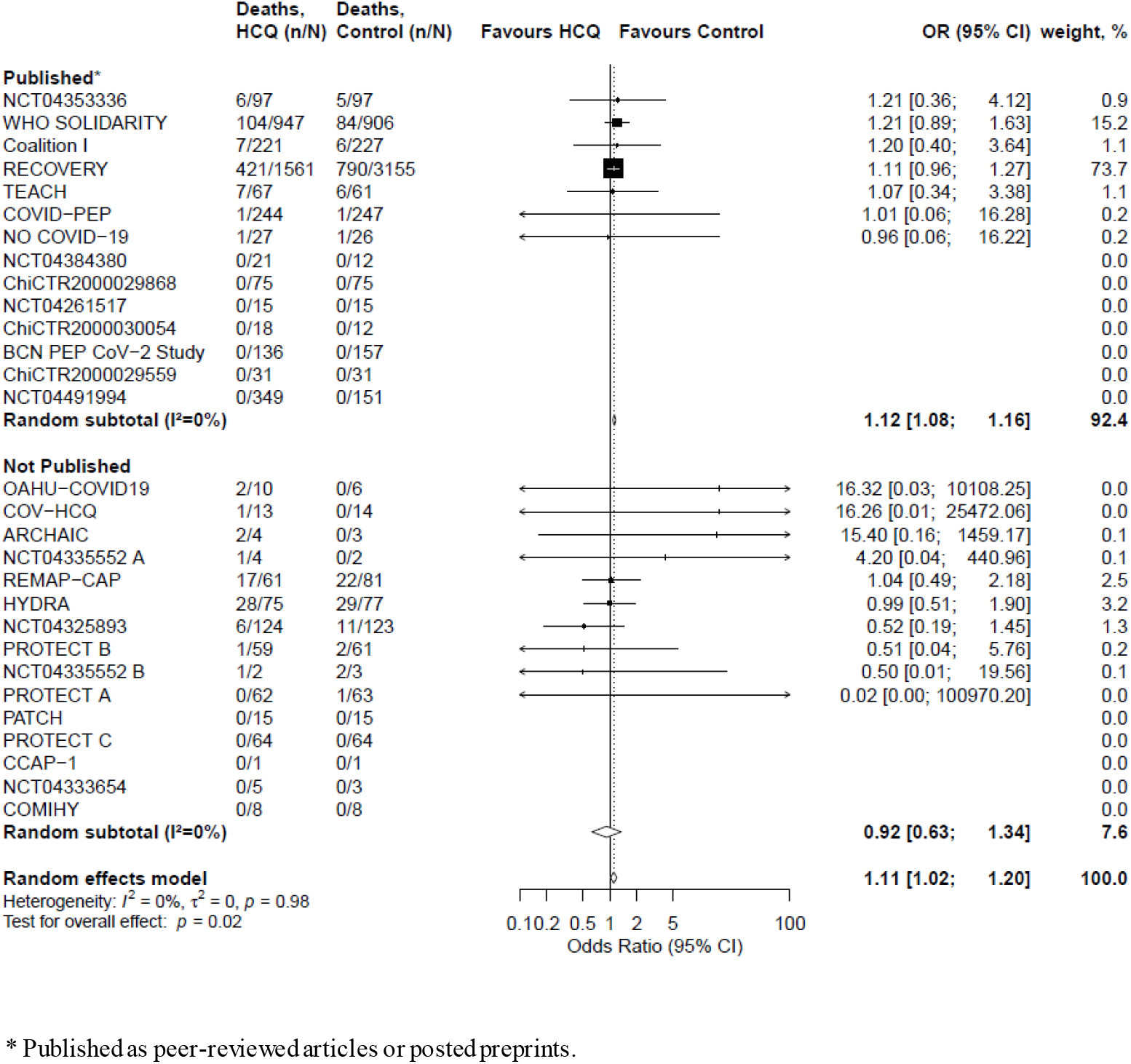
Random effects meta-analysis for mortality for treatment of COVID-19 with Hydroxychloroquine, trials are stratMortality outcomes with hydroxychloroquine and chloroquineified by publication status.

Regarding CQ, in the four included trials, 18 of 160 (11%) patients treated with CQ died and 12 of 147 patients (8%) in the control groups died. The combined OR was 1.77 (95% CI: 0.15 to 21.13, p = 0.21), with low heterogeneity (I^2^ = 0%) (Figure 2B). In two of four trials including a total of 217 patients, there were zero deaths in both arms.

**Figure 2B.**
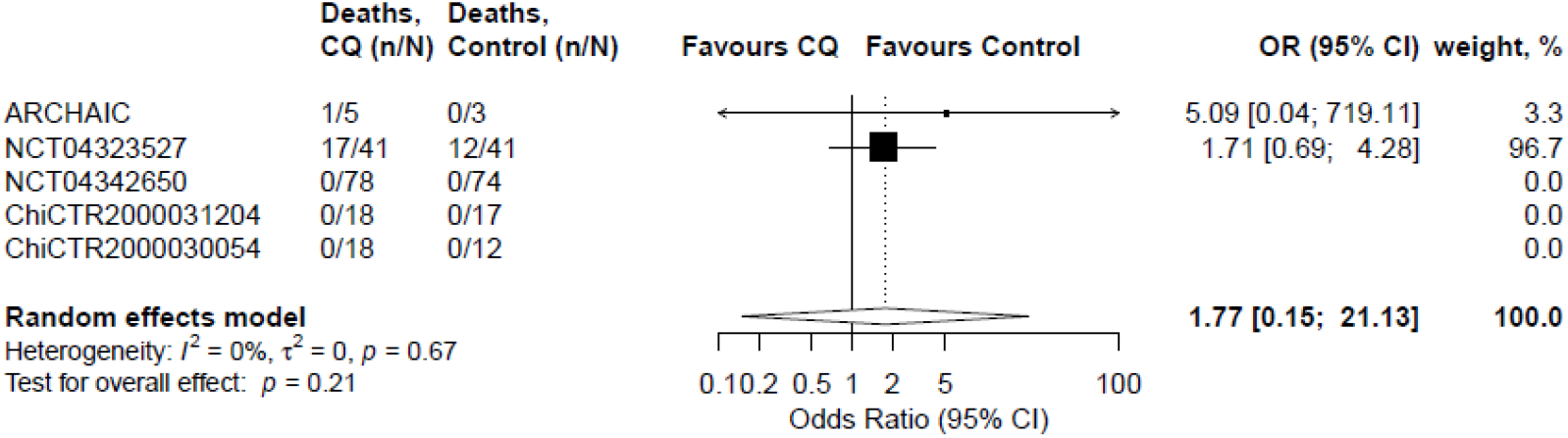
Random effects meta-analysis for mortality for treatment of COVID-19 with Chloroquine. The x-axis scales differ for reasons of readability.

The available evidence in this study is the result of publications, preprints or personal communication accrued from April 10, 2020 to October 15, 2020 as shown in the cumulative meta-analyses (Figure 3A-C).

**Figure 3A.**
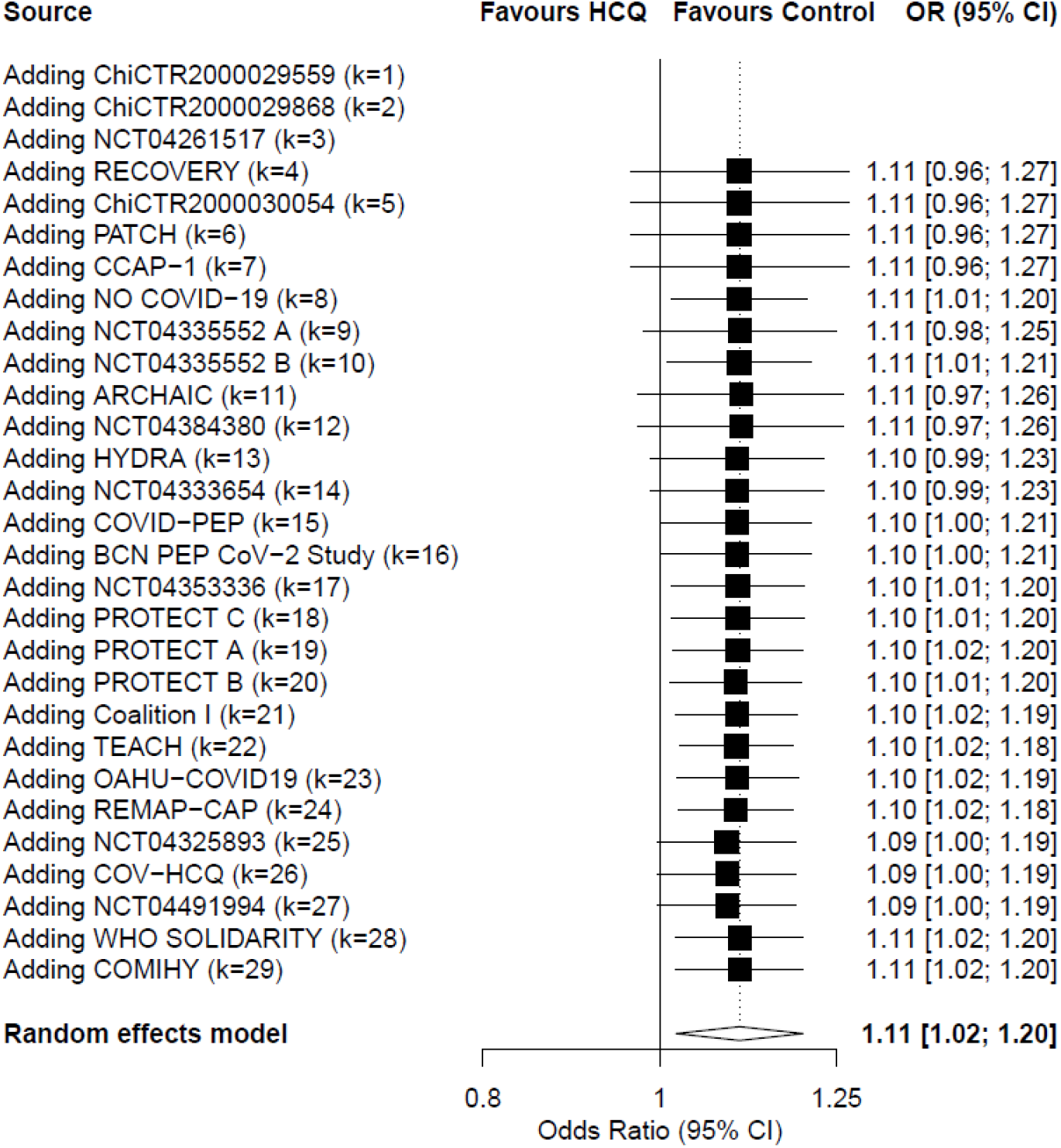
Cumulative meta-analysis for mortality for treatment of COVID-19 with Hydroxychloroquine.

**Figure 3B.**
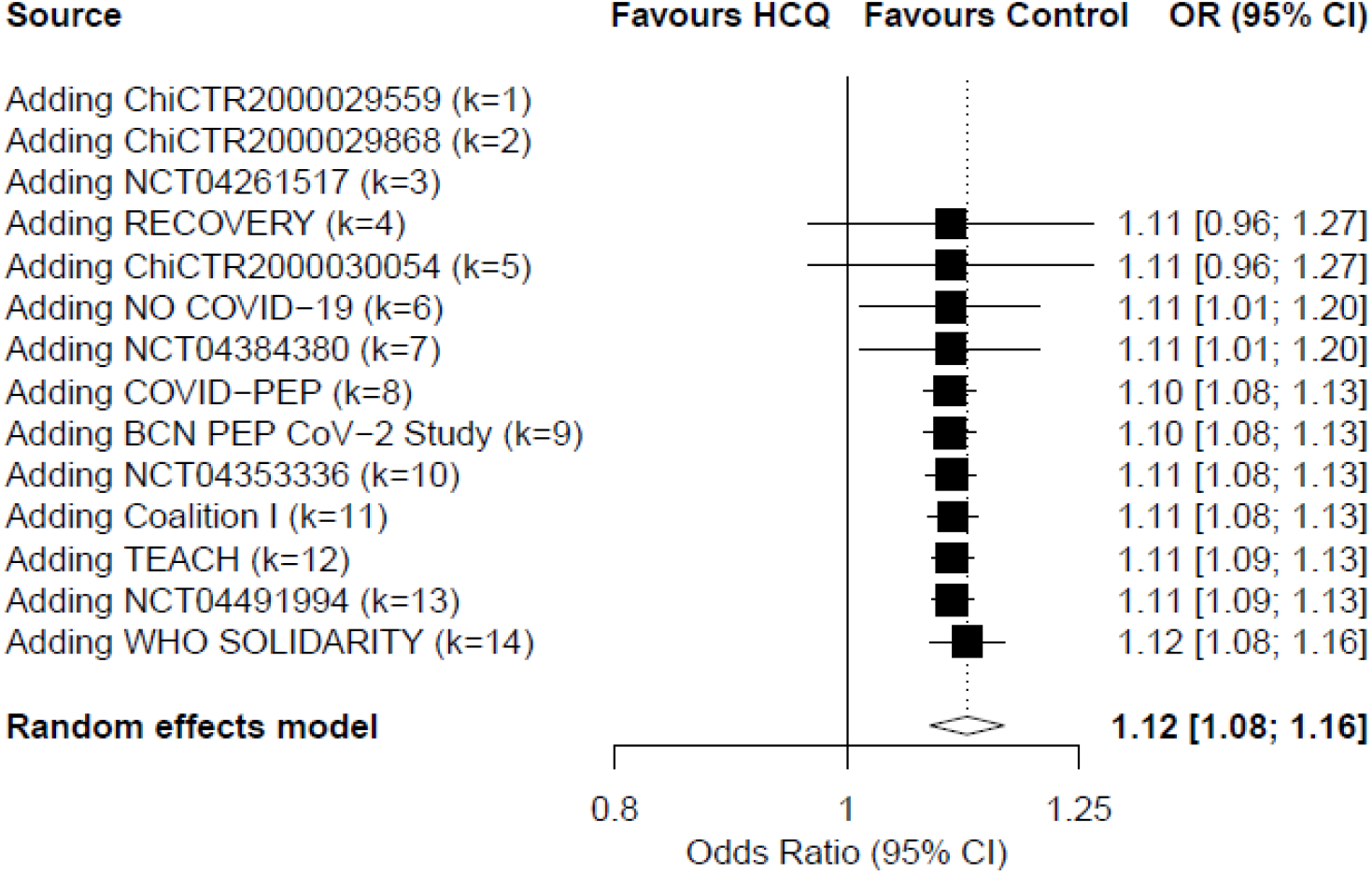
Cumulative meta-analysis for mortality for treatment of COVID-19 with Hydroxychloroquine (publications and preprints only)

**Figure 3C.**
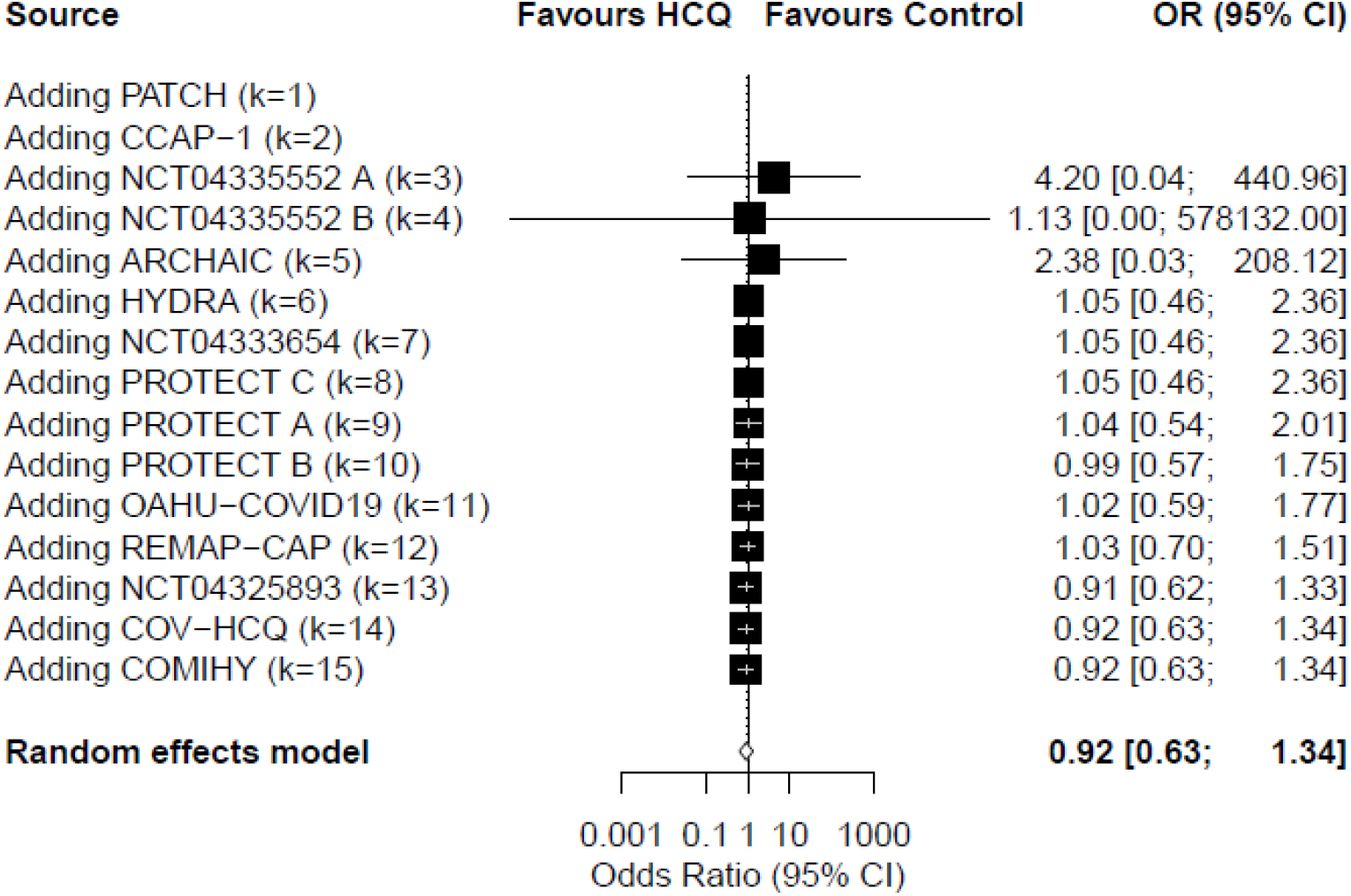
Cumulative meta-analysis for mortality for treatment of COVID-19 with Hydroxychloroquine (unpublished data only) The x-axis scales differ for reasons of readability.

For HCQ, none of the exploratory subgroup analyses showed an effect modification (Supplement Table S2A). When only including published information (publications and preprints, excluding unpublished trials), there was an increase in mortality among patients treated with HCQ (OR 1.12, 95% CI 1.08 to 1.16), while among the unpublished trials there was no such sign of increased mortality(OR 0.92, 95% CI 0.63 to 1.34, *p* for interaction = 0.23). We conducted no subgroup analyses for CQ, as there were only two trials with events. In the sensitivity analyses employing different meta-analytical approaches (Supplement Table S2B and Figures S1A-C), results were compatible with the main analysis.

## Discussion

This collaborative meta-analysis of 28 published or unpublished RCTs, including 10,319 patients, shows that treatment with HCQ was associated with increased mortality in COVID-19 patients, and there was no benefit from treatment with CQ. No differences were seen across subgroup analyses on patient setting, diagnosis confirmation, control type, publication status or dose and the between-study heterogeneity was low. For CQ, the number of studies was too small to draw clear conclusions.

This meta-analysis offers useful insights for a challenging health situation. Hundreds of thousands of patients have received HCQ and CQ outside of clinical trials without evidence of their beneficial effects. Public interest is unprecedented, with weak early evidence supporting HCQ ‘s merits being widely discussed in some media and social networks - despite the unfavorable results by a very large RCT. Numerous clinical studies have been investigating HCQ and CQ almost simultaneously. Although several systematic reviews and meta-analyses are already available, they only consider the small handful of RCTs being already published (which were all included here).^36–40^ While data sharing has been rather limited to-date in biomedical research, such openness can be transformative in generating knowledge. This pandemic has brought together a collaboration of clinical trialists agreeing to share their data, which allows this study to not only summarize the existing evidence, but also illustrate the accumulation of evidence that would otherwise not be available.

For HCQ, evidence is dominated by the RECOVERY trial,^13^which indicated no mortality benefit for treated COVID-19 patients, together with longer hospitalization and higher risk of progression to invasive mechanical ventilation and/or death. Similarly, the WHO SOLIDARITY trial indicated no mortality benefit.^35^ The RECOVERY and WHO SOLIDARITY trials used HCQ in comparatively higher doses than all other trials except REMAP-CAP. There was no evidence for an effect modification by dose (p for interaction = 0.29), and the combined effect of all the trials with lower dose did not indicate a benefit of HCQ but tended to a null effect (OR 0.97; 0.73 to 1.30) with wide CIs, compatible with the main effect estimate.

Although the published trials resulted in a conclusive treatment estimate, the unpublished trials tend towards a null effect. The tendency of published trials to report larger effect sizes than unpublished trials is well-documented and constitute one of the reporting biases that are discernable only when a body of studies are considered together.^41^Null results are less expected to be rapidly disseminated, especially if the trial is small. Of note, RECOVERY results showing dexamethasone benefits have been published more rapidly ^3^ than the unfavorable HCQ results.^13^This paper offers the most comprehensive summary on HCQ and mortality in COVID-19 to date.

This meta-analysis does not address prophylactic use nor other outcomes besides mortality. All but three trials excluded children and the majority excluded pregnant or breastfeeding women; generalizability remains unclear for those populations. Among the five studies on outpatients, there were three deaths, two occurring in the one trial of 491 relatively young patients with few comorbidities,^25^and one occurring in a small trial with 27 patients. For outpatients that are elderly or have comorbidities, evidence is sparse. Most of the 28 trials excluded persons with comorbid conditions carrying higher risk of adverse events from HCQ or CQ.^23–25^No evidence is in the pipeline for these groups, which echoes clinical reasoning being reluctant to expose them to risk.

Twenty-three percent of the potentially eligible trials were listed as discontinued, mostly because of fewer patients than expected. Among 28 included RCTs, only two had reached their target sample size at the time of censoring for this meta-analysis. As previously discussed,^42^ most trials on HCQ and CQ in COVID-19 are small, reflecting both the strong motivation for individual efforts and underscoring the need for readily available research infrastructure to merge small-scale initiatives.^4^ Especially in the context of recruitment challenges, we encourage other researchers to form collaborations and combine trial results.^43^

Our analysis has some limitations. First, although we adopted a comprehensive, systematic search strategy, our real-time initiative differs from traditional systematic reviews. We focused on collecting unpublished information, aiming to rapidly secure as much trial evidence as possible. We did not review individual trials, nor did we stratify results according to patient characteristics. Such analyses are planned in future publications using in-depth details disclosed in individual trial publications to come.^44–46^ The exploratory subgroup analyses did not support the hypothesis that blinding/use of placebo is associated with the observed effect (the test for an interaction gives p=0.15 and the OR is 0.88 with wide CIs [0.55 to 1.41], compatible with the overall effect); moreover, attrition was negligible (median 0%, IQR 0% to 0%; range 0 to 19.5%). A meta-epidemiological study shows little evidence that mortality results would be affected by lack of blinding, or problems in randomization and allocation concealment, in contrast to less objective outcomes.^47^Second, a majority of the potentially eligible trials were not available. Despite going far beyond the standard review of published evidence, we expect additional results from future trials on CQ to narrow the uncertainty of the treatment effect and possibly reveal benefits or harms not discernible based on the current evidence. We plan to perform an update when substantial additional evidence becomes available. Finally, although sensitivity analyses addressing model specifications were compatible with the main analysis, one combination (HKSJ model with SJ tau-squared estimator) yielded substantially wider confidence intervals. This combination gave disproportionately low weight to RECOVERY (15%) and we consider the main model (HKSJ with PM tau-squared estimator) to be more valid in this situation.

Treatment with HCQ for COVID-19 was associated with increased mortality, and there was no benefit from CQ based on currently available randomized trial data. Medical professionals around the globe are encouraged to inform patients about this evidence.

## Data Availability

All data generated or analysed during this study are included in this published article and its supplementary information files.

## Declarations

### Authors’ contributions

Lars G. Hemkens, Cathrine Axfors and Andreas M. Schmitt had full access to all data in this study and take responsibility for the integrity of the data and the accuracy of the data analysis.

*Concept and design*: Lars G. Hemkens, John P. A. Ioannidis, Cathrine Axfors, Andreas M. Schmitt, Steven N. Goodman, David Moher

*Acquisition, analysis, or interpretation of data*: All authors

*Drafting of the manuscript:* Cathrine Axfors, Andreas M. Schmitt, Lars G. Hemkens

*Critical revision of the manuscript for important intellectual content*: All authors

*Statistical analysis:* Andreas M. Schmitt, Lars G. Hemkens, John P. A. Ioannidis, Steven N. Goodman, Perrine Janiaud

*Approval of the final manuscript*: All authors

*Obtained funding*: Lars G. Hemkens, Cathrine Axfors, John P. A. Ioannidis

*Administrative, technical, or material support*: Cathrine Axfors, Andreas M. Schmitt, Lars G. Hemkens, Perrine Janiaud, Janneke van ‘t Hooft

*Supervision*: Lars G. Hemkens

## Acknowledgements

We wish to express our heartfelt gratitude to all patients volunteering for the trials involved. We furthermore thank Wenyan Ma and Benjamin Kasenda (University Hospital Basel, University of Basel) for kindly translating the emails to Chinese investigators. For valuable contributions to individual trials in this collaborative group, we sincerely thank: Hannah Jin, Monica Feeley, Bruce Bausk, Jessica Cauley, Jane Kleinjan, Jon Gothing, Naeema Bangash, Heather Wroe, Claire Bigogne, Christelle Castell, Annelies Mottart, Lisette Cortenraad, Judith Medema-Muller, Katia Handelberg, Khalid Benhammou, Shaheen Kumar, Sophie Gribomont, Kim Kuehne, Cathia Markina, Julien Labeirie, Julie Pencole, Eva Crispyn, Cecile Le Breton, Kelli Horn, Tina Patel, Benjamin Harrois, Isabelle Collin, Vetheeswar Manivannan, Irma Slomp, Frederick Becue, Isabelle Godefroy, Lynne Guo, Lene Kollmorgen, Toluwalope Cole, Catherine Chene, Praveena Deenumsetti, Anne Doisy, Ariane Vialfont, Melissa Charbit, Christine Shu, Stephane Kirkesseli, Howard Surks, Magalie De Meyer, Edel Hendrickx, and Paul Deutsch (Sanofi trial); Ellie Carmody, Märtin Backer, Jaishvi Eapen, Jack A. DeHovitz, Prithiv J. Prasad, Yi Li, Camila Delgado, Morris Jrada, Gabriel A. Robbins, Brooklyn Henderson, Alexander Hrycko, Dinuli Delpachitra, Vanessa Raabe, Jonathan S. Austrian, and Yanina Dubrovskaya (TEACH trial); Farah Al-Beidh, Djillali Annane, Kenneth Baillie, Abigail Beane, Richard Beasley, Zahra Bhimani, Marc Bonten, Charlotte Bradbury, Frank Brunkhorst, Meredith Buxton, Allen Cheng, Menno de Jong, Eamon Duffy, Lise Estcourt, Rob Fowler, Timothy Girard, Herman Goossens, Cameron Green, Rashan Haniffa, Christopher Horvat, David Huang, Francois Lamontagne, Patrick Lawler, Kelsey Linstrum, Edward Litton, John Marshall, Daniel McAuley, Shay McGuinness, Stephanie Montgomery, Paul Mouncey, Katrina Orr, Rachael Parke, Jane Parker, Asad Patanwala, Kathryn Rowan, Marlene Santos, Christopher Seymour, Steven Tong, Anne Turner, Timothy Uyeki, Wilma van Bentum-Puijk, Frank van de Veerdonk, and Ryan Zarychanski (REMAP-CAP trial); Jan-Erik Berdal, Arne Eskesen, Dag Kvale, Inge Christoffer Olsen, Corina Silvia Rueegg, Anbjørg Rangberg, Christine Monceyron Jonassen, and Torbjørn Omland (NO COVID-19 Trial). Support for title page creation and format was provided by AuthorArranger, a tool developed at the National Cancer Institute.

## Supplements to “Mortality outcomes with hydroxychloroquine and chloroquine in COVID-19: an international collaborative meta-analysis of randomized trials”

1. **Search strategy**
2. **Template for invitation email**
3. **Table S1. Group-level characteristics of unavailable trials**
4. **Table S2A. Subgroup analyses**
5. **Table S2B. Sensitivity analyses**
6. **Figures S1A-C. Forest plots for sensitivity analyses**

## Supplement 1. Search strategy

The COVID-evidence database includes trials registered on ClinicalTrials.gov or the WHO International Clinical Trials Registry Platform up to June 11, 2020, as well as trials published on the following sources up to April 9, 2020: PubMed, medrXiv, biorXiv, the WHO COVID-19 literature database, and a listing of all trials with ethical approval in Switzerland (for details please see the COVID-evidence study protocol on the Open Science Framework: http://dx.doi.org/10.17605/OSF.IO/GEHFX).

This Supplement describes the search strategy used to complement the COVID-evidence database with trials registered or published after April 9, 2020.

PubMed and the Cochrane COVID-19 trial registry were searched from inception to June 11, 2020. Search terms included extensive controlled vocabulary and Medical Subject Headings (MeSH). Search terms were the following: corona[ti] OR covid*[ti] OR sars[ti] OR severe acute respiratory syndrome[ti] OR ncov*[ti] OR “severe acute respiratory syndrome coronavirus 2” [Supplementary Concept] OR “COVID-19” [Supplementary Concept] OR (wuhan[tiab] AND coronavirus[tiab]) OR (wuhan[tiab] AND pneumonia virus[tiab]) OR COVID19[tiab] OR COVID-19[tiab] OR coronavirus 2019[tiab] OR SARS-CoV-2[tiab] OR SARS2[tiab] OR SARS-2[tiab] OR “severe acute respiratory syndrome 2”[tiab] OR 2019-nCoV[tiab] OR (novel coronavirus[tiab] AND 2019[tiab]) NOT (animals[mesh] NOT humans[mesh]) AND (“2019/12/01”[EDAT] : “3000/12/31”[EDAT]) AND (((hydroxychloroquine[MeSH Terms]) OR (chloroquine[MeSH Terms])) OR (hydroxychloroquine[Title/Abstract])) OR (chloroquine[Title/Abstract]) AND (randomized controlled trial[pt] OR controlled clinical trial[pt] OR randomized[tiab] OR placebo[tiab] OR clinical trials as topic[mesh:noexp] OR randomly[tiab] OR trial[ti] NOT (animals[mh] NOT humans [mh]))

The search was updated on October 16, 2020.

## Supplement 2. Template for invitation email

Subject: Invitation to co-author a large-scale international collaborative meta-analysis on mortality in COVID-19 trials

Dear Dr. **<last name>**,

We are currently conducting a large-scale international collaborative meta-analysis on mortality in all ongoing or completed randomized clinical trials evaluating hydroxychloroquine or chloroquine for COVID-19. We are inviting all research groups worldwide testing these drugs to provide urgently needed evidence. We have no commercial interest with this work and aim to rapidly publish the results in a peer-reviewed journal. **Your core team is invited to co-author the publication**.

We use our COVID-evidence database (www.covid-evidence.org) for this work. COVID-evidence is supported by the Swiss National Science Foundation (Project ID 196190) and a large collaboration of researchers from Switzerland, the US, China, Canada, UK, France, Germany, Austria, Sweden, Netherlands, and other countries.

Our registered protocol can be found **attached** as well as registered on the Open Science Framework: **[link]**. Trials that are eligible for this project can be found at the end of the protocol.

Your study **<url>** is of high importance for this project. We would like to ask you how many patients died in your trial (see short questions below). We will use standard methods of meta-analysis and focus only on group-level (aggregated) mortality (no individual patient data needed). We will not do an in-depth review of the included trials as we aim to rapidly provide results, ideally from all trials worldwide. We describe the details of the study in the attached protocol.

**For the meta-analysis, we kindly ask you to answer the following questions before July 7. If you are interested in collaborating, please let us know of your interest as soon as possible**.

---

**Question 1**: Could you please confirm that these criteria apply to your trial?

a. The trial is randomized and started enrollment before June 1, 2020
b. The trial has at least one group of patients who receive hydroxychloroquine or chloroquine
c. The trial has at least one control group that does not receive hydroxychloroquine or chloroquine

**Question 2**: For each of your study arms,

a. What intervention did this group receive?
b. How many patients were randomized to this group?
c. Of these patients, how many have died?
d. Of these patients, for how many it is unknown if they are dead or alive?

---

Please note that we are interested in these raw numbers regardless of the results of any statistical test. The numbers will be used to finalize the manuscript described in our registered protocol (attached).

We strive for a rapidly available, maximally informative publication with full transparency on methods. **With this publication we aim to make sure that all clinical trial data hitherto collected (unpublished or published) will be of use, regardless of whether the target sample size of each trial was reached or not. We invite your core investigator team as co-authors**. The manuscript will be shared with all co-authors for comments and the finalized manuscript will be uploaded as a preprint at medrXiv in parallel with submission to a peer-reviewed medical journal such as JAMA or the BMJ.

Thank you for considering our request! We kindly ask for your answer **before July 7**. If you are interested in collaborating, but are uncertain whether the data may be shared before July 7, please respond as soon as possible. Should you have any questions or comments, please let us know.

Best regards,

Cathrine Axfors, Andreas Schmitt, David Moher, Steve Goodman, John Ioannidis and Lars Hemkens for the COVID-evidence team

www.covid-evidence.org

**Table S1.**
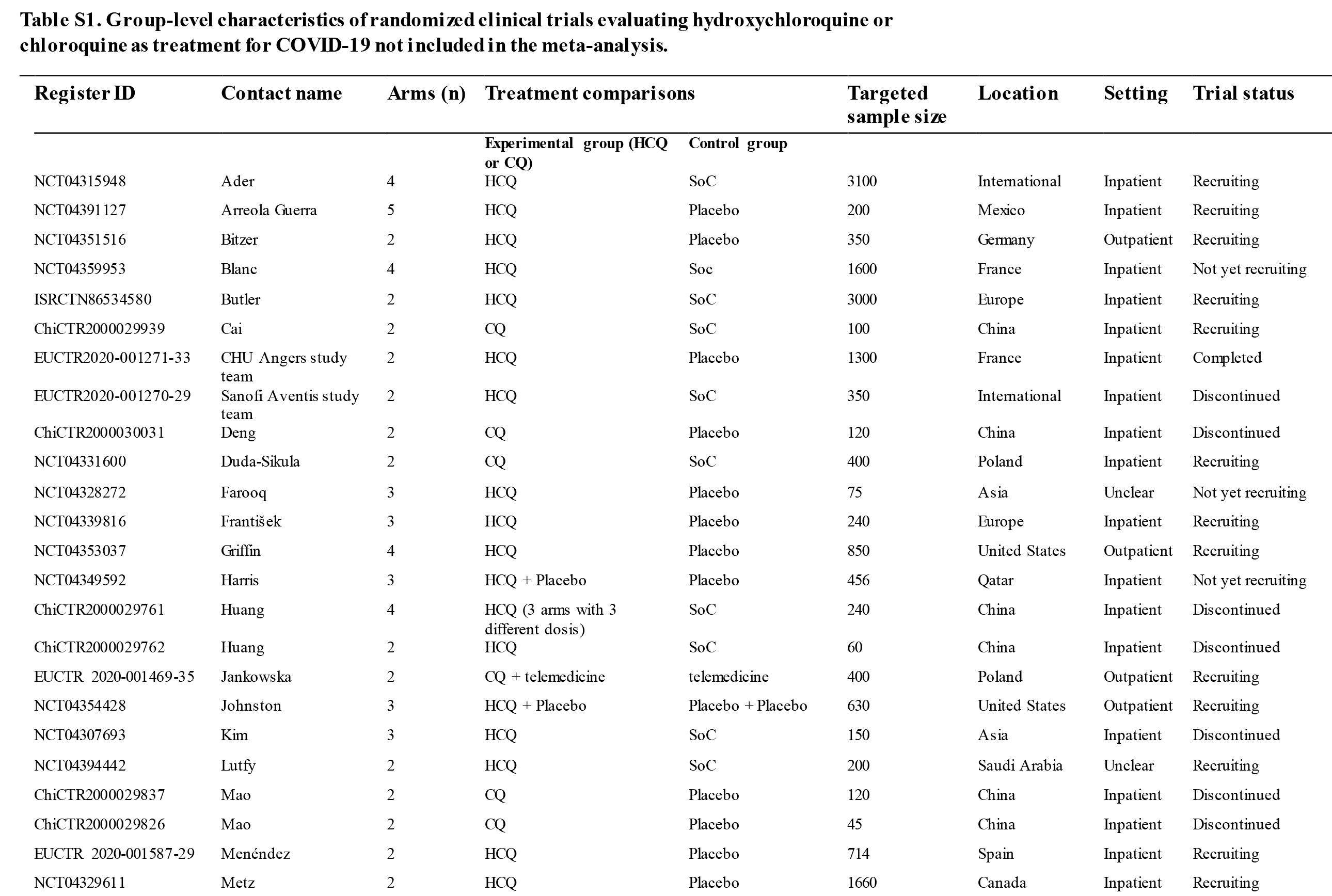
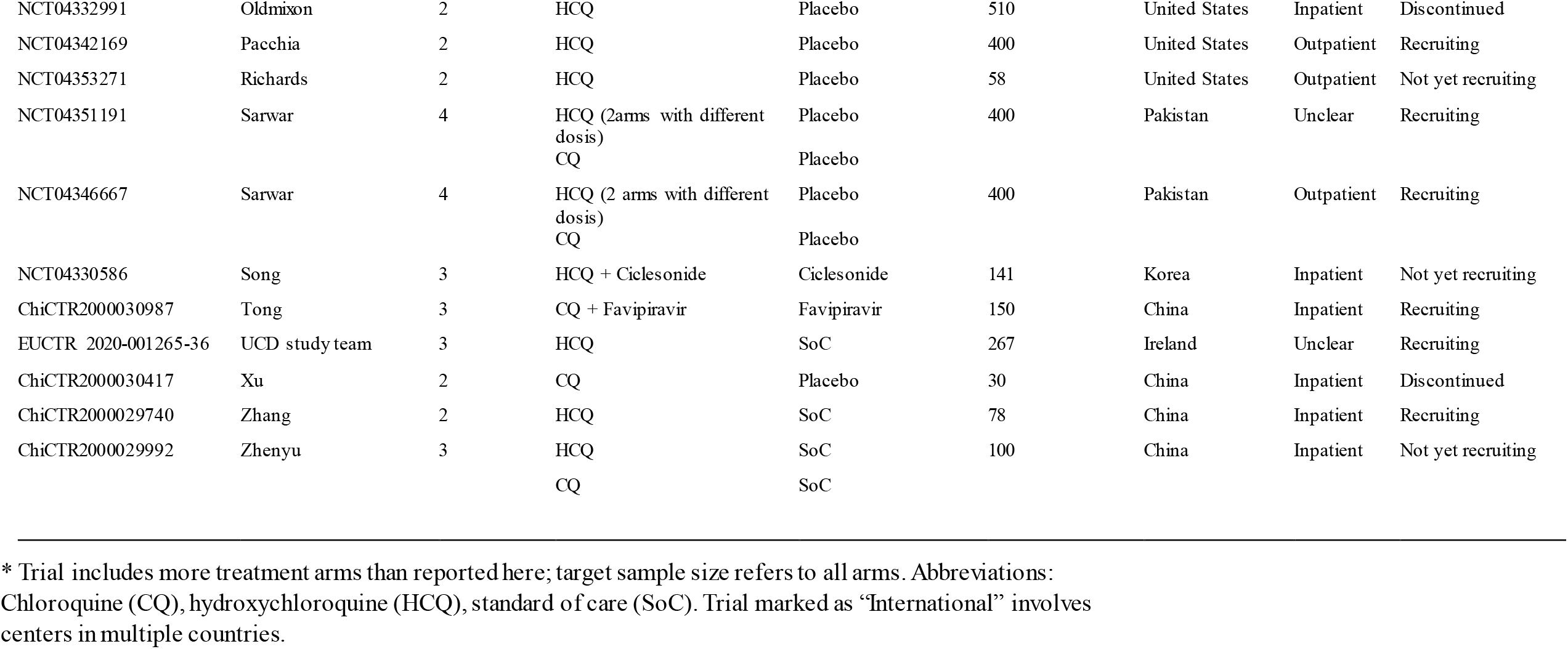
Group-level characteristics of randomized clinical trials evaluating hydroxychloroquine or chloroquine as treatment for COVID-19 not included in the meta-analysis.

| Register ID | Contact name | Arms (n) | Treatment comparisons |  | Targeted sample size | Location | Setting | Trial status |
| --- | --- | --- | --- | --- | --- | --- | --- | --- |
|  |  |  | Experimental group (HCQ or CQ) | Control group |  |  |  |  |
| NCT04315948 | Ader | 4 | HCQ | SoC | 3100 | International | Inpatient | Recruiting |
| NCT04391127 | Arreola Guerra | 5 | HCQ | Placebo | 200 | Mexico | Inpatient | Recruiting |
| NCT04351516 | Bitzer | 2 | HCQ | Placebo | 350 | Germany | Outpatient | Recruiting |
| NCT04359953 | Blanc | 4 | HCQ | Soc | 1600 | France | Inpatient | Not yet recruiting |
| ISRCTN86534580 | Butler | 2 | HCQ | SoC | 3000 | Europe | Inpatient | Recruiting |
| ChiCTR2000029939 | Cai | 2 | CQ | SoC | 100 | China | Inpatient | Recruiting |
| EUCTR2020-001271-33 | CHU Angers study team | 2 | HCQ | Placebo | 1300 | France | Inpatient | Completed |
| EUCTR2020-001270-29 | Sanofi Aventis study team | 2 | HCQ | SoC | 350 | International | Inpatient | Discontinued |
| ChiCTR2000030031 | Deng | 2 | CQ | Placebo | 120 | China | Inpatient | Discontinued |
| NCT04331600 | Duda-Sikula | 2 | CQ | SoC | 400 | Poland | Inpatient | Recruiting |
| NCT04328272 | Farooq | 3 | HCQ | Placebo | 75 | Asia | Unclear | Not yet recruiting |
| NCT04339816 | František | 3 | HCQ | Placebo | 240 | Europe | Inpatient | Recruiting |
| NCT04353037 | Griffin | 4 | HCQ | Placebo | 850 | United States | Outpatient | Recruiting |
| NCT04349592 | Harris | 3 | HCQ + Placebo | Placebo | 456 | Qatar | Inpatient | Not yet recruiting |
| ChiCTR2000029761 | Huang | 4 | HCQ (3 arms with 3 different dosis) | SoC | 240 | China | Inpatient | Discontinued |
| ChiCTR2000029762 | Huang | 2 | HCQ | SoC | 60 | China | Inpatient | Discontinued |
| EUCTR 2020-001469-35 | Jankowska | 2 | CQ + telemedicine | telemedicine | 400 | Poland | Outpatient | Recruiting |
| NCT04354428 | Johnston | 3 | HCQ + Placebo | Placebo + Placebo | 630 | United States | Outpatient | Recruiting |
| NCT04307693 | Kim | 3 | HCQ | SoC | 150 | Asia | Inpatient | Discontinued |
| NCT04394442 | Lutfy | 2 | HCQ | SoC | 200 | Saudi Arabia | Unclear | Recruiting |
| ChiCTR2000029837 | Mao | 2 | CQ | Placebo | 120 | China | Inpatient | Discontinued |
| ChiCTR2000029826 | Mao | 2 | CQ | Placebo | 45 | China | Inpatient | Discontinued |
| EUCTR 2020-001587-29 | Menéndez | 2 | HCQ | Placebo | 714 | Spain | Inpatient | Recruiting |
| NCT04329611 | Metz | 2 | HCQ | Placebo | 1660 | Canada | Inpatient | Recruiting |
| NCT04332991 | Oldmixon | 2 | HCQ | Placebo | 510 | United States | Inpatient | Discontinued |
| NCT04342169 | Pacchia | 2 | HCQ | Placebo | 400 | United States | Outpatient | Recruiting |
| NCT04353271 | Richards | 2 | HCQ | Placebo | 58 | United States | Outpatient | Not yet recruiting |
| NCT04351191 | Sarwar | 4 | HCQ (2arms with different<br>dosis)<br>CQ | Placebo<br>Placebo | 400 | Pakistan | Unclear | Recruiting |
| NCT04346667 | Sarwar | 4 | HCQ (2 arms with different<br>dosis)<br>CQ | Placebo<br>Placebo | 400 | Pakistan | Outpatient | Recruiting |
| NCT04330586 | Song | 3 | HCQ + Ciclesonide | Ciclesonide | 141 | Korea | Inpatient | Not yet recruiting |
| ChiCTR2000030987 | Tong | 3 | CQ + Favipiravir | Favipiravir | 150 | China | Inpatient | Recruiting |
| EUCTR 2020-001265-36 | UCD study team | 3 | HCQ | SoC | 267 | Ireland | Unclear | Recruiting |
| ChiCTR2000030417 | Xu | 2 | CQ | Placebo | 30 | China | Inpatient | Discontinued |
| ChiCTR2000029740 | Zhang | 2 | HCQ | SoC | 78 | China | Inpatient | Recruiting |
| ChiCTR2000029992 | Zhenyu | 3 | HCQ<br>CQ | SoC<br>SoC | 100 | China | Inpatient | Not yet recruiting |

\* Trial includes more treatment arms than reported here; target sample size refers to all arms. Abbreviations: Chloroquine (CQ), hydroxychloroquine (HCQ), standard of care (SoC). Trial marked as “International” involves centers in multiple countries.

**Table S2A.** Subgroup analyses for random-effects meta-analysis on mortality for treatment of COVID-19 with Hydroxychloroquine.

| Subgroup | Trials (n) | Patients (n) | OR (95% CI) | P value, test for interaction |
| --- | --- | --- | --- | --- |
| **Setting** |  |  |  |  |
| ICU | 1 | 142 | 1.04 (0.49, 2.18) | 0.98 |
| Inpatient | 21 | 9062 | 1.11 (1.02, 1.21) |  |
| Outpatient | 4 | 808 | 1.01 (0.06, 16.28) |  |
| **Published** |  |  |  |  |
| Yes | 14 | 8981 | 1.12 (1.08, 1.16) | 0.23 |
| No | 12 | 1031 | 0.92 (0.63, 1.34) |  |
| **Control** |  |  |  |  |
| Standard of care | 17 | 8911 | 1.12 (1.04, 1.21) | 0.15 |
| Placebo | 9 | 1101 | 0.88 (0.55, 1.41) |  |
| **COVID-19 diagnostic confirmation** |  |  |  |  |
| Confirmed | 22 | 4215 | 1.11 (0.91, 1.36) | 0.94 |
| Confirmed or suspected | 4 | 5797 | 1.1 (1.07, 1.14) |  |
| **Dose\*** |  |  |  |  |
| High | 3 | 6711 | 1.12 (1.01, 1.25) | 0.29 |
| Low | 23 | 3301 | 0.97 (0.73, 1.30) |  |

\* High dose: $\geq 1600$ mg on day 1 and $\geq 800$ mg from day 2. Low dose: $< 1600$ mg on day 1 or $< 800$ from day 2.

**Table S2B.**
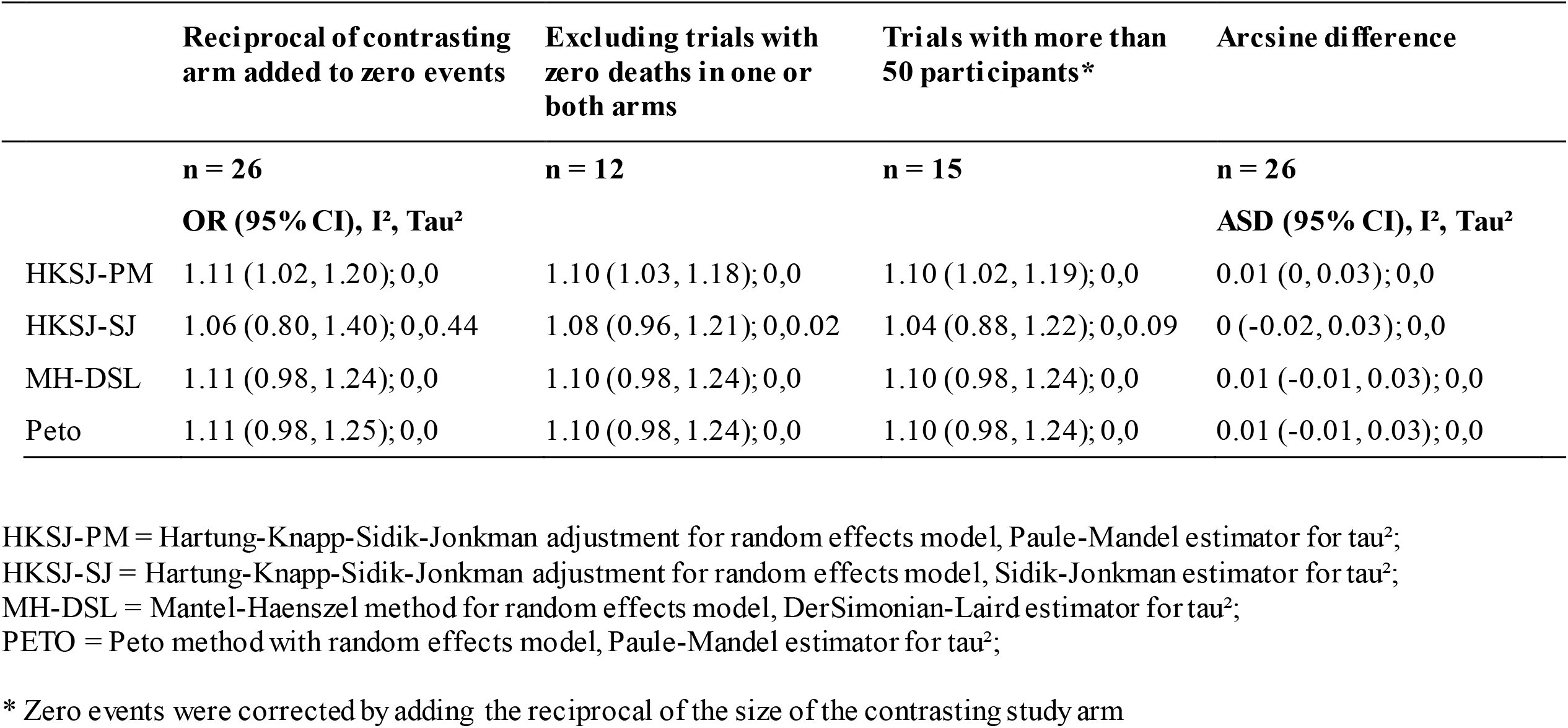
Sensitivity analysis for meta-analysis for mortality for treatment of COVID-19 with Hydroxychloroquine using different methods of combination.

**Figure S1A.**
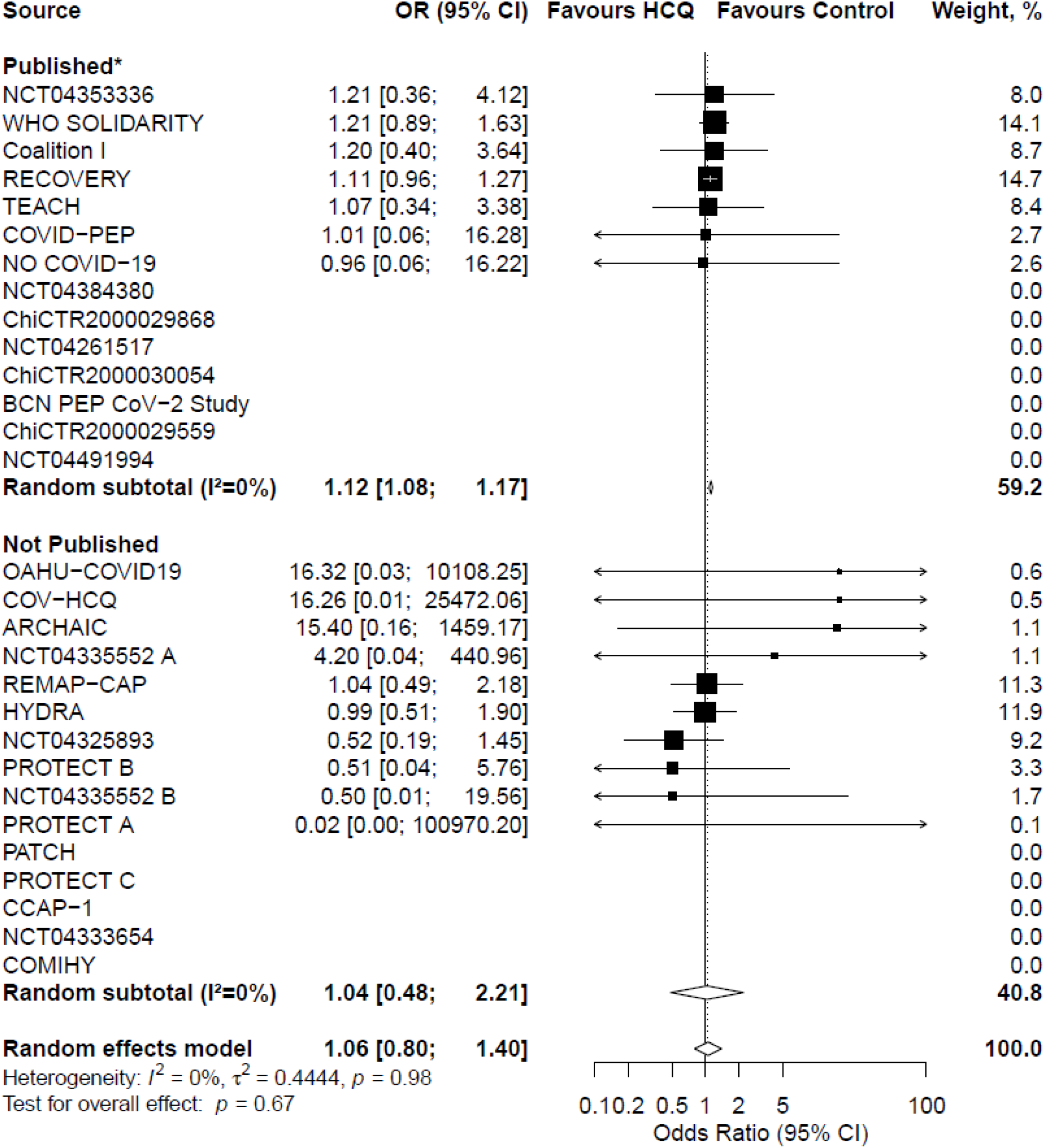
Forest plot of HKSJ-SJ model.

Zero events were corrected by adding the reciprocal of the size of the contrasting study arm.

**Figure S1B.**
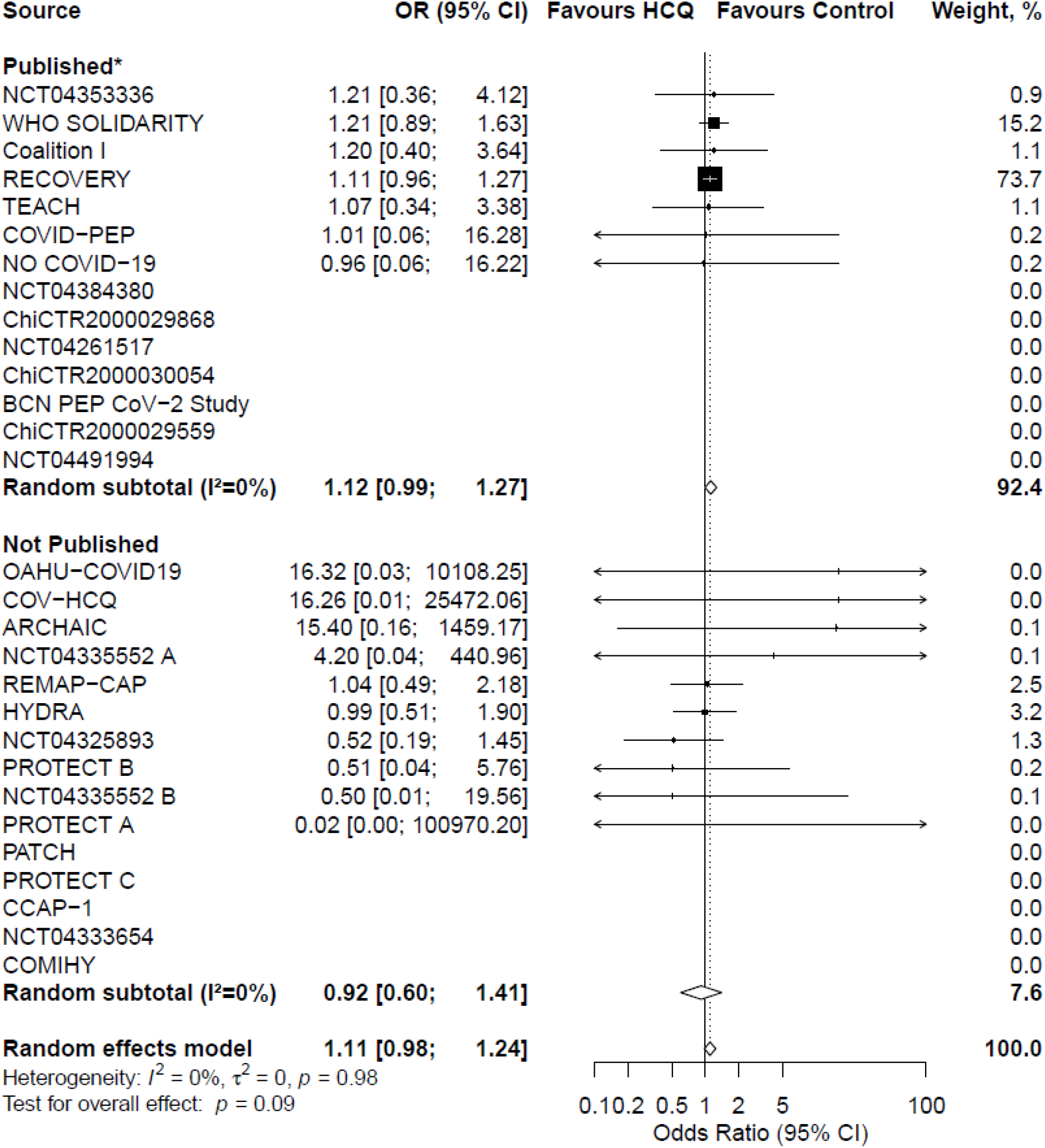
Forest plot of MH-DSL model. Zero events were corrected by adding the reciprocal of the size of the contrasting study arm.

**Figure S1C.**
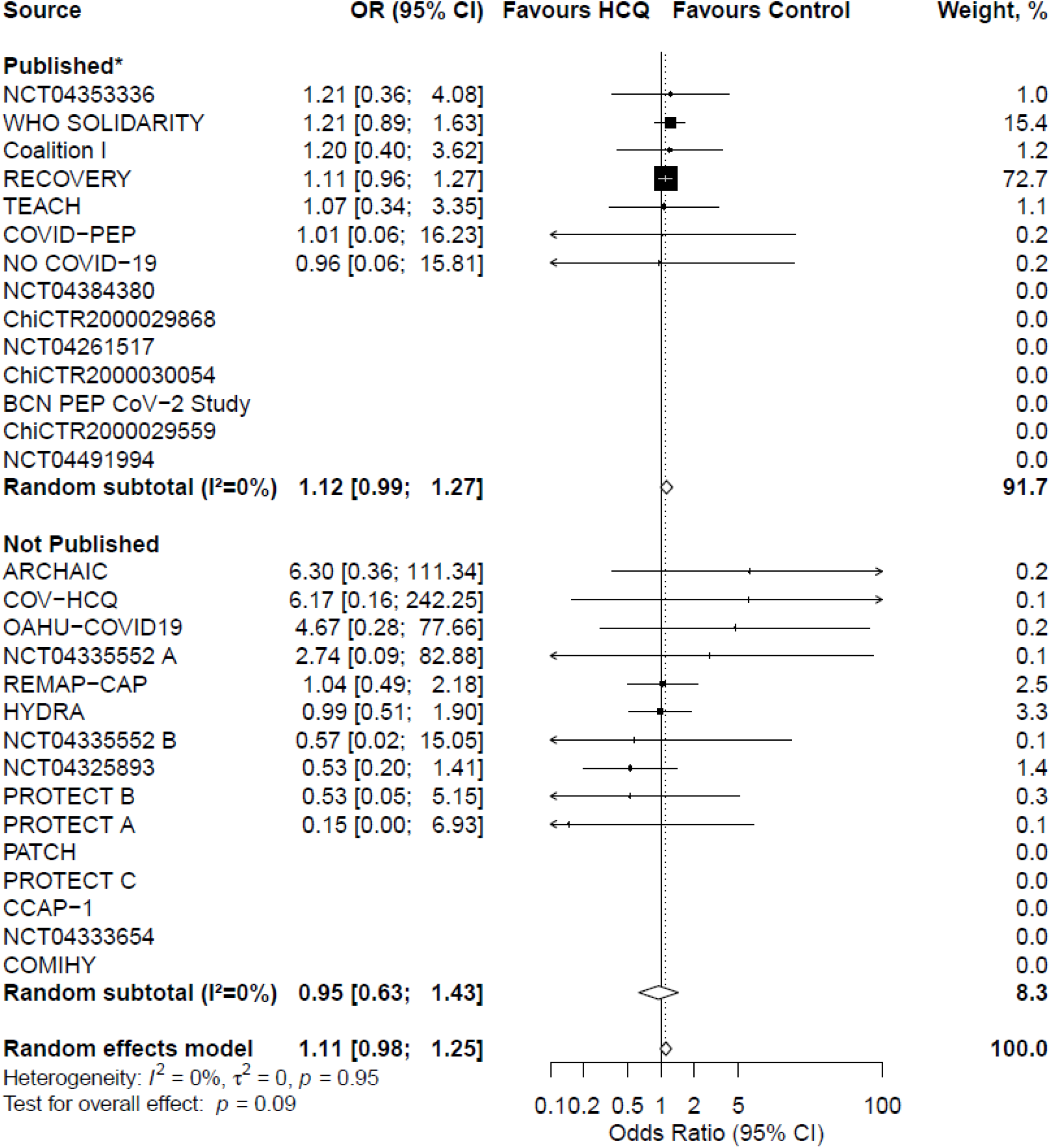
Forest plot of Peto model. Zero events were corrected by adding the reciprocal of the size of the contrasting study arm.

